# The mouth of America: the oral microbiome profile of the US population

**DOI:** 10.1101/2024.12.03.24318415

**Authors:** Anil K. Chaturvedi, Emily Vogtmann, Jianxin Shi, Yukiko Yano, Martin J. Blaser, Nicholas A. Bokulich, J. Gregory Caporaso, Maura L. Gillison, Barry I. Graubard, Xing Hua, Autumn G. Hullings, Lisa Kahle, Rob Knight, Shilan Li, Jody McLean, Vaishnavi Purandare, Yunhu Wan, Neal D. Freedman, Christian C. Abnet

**Author notes:** **Correspondence:** Christian C. Abnet, PhD Director, Metabolic Epidemiology Branch Division of Cancer Epidemiology and Genetics, National Cancer Institute 9609 Medical Center Drive, Rm. 6E410 Rockville, MD. 20852. These authors contributed equally to the work (Anil K. Chaturvedi, Emily Vogtmann, Neal D. Freedman, Christian C. Abnet).

## Abstract

**Importance:** The oral microbiome is increasingly recognized to play key roles in human health and disease; yet, population-representative characterizations are lacking.

**Objective:** Characterize the composition, diversity, and correlates of the oral microbiome among US adults.

**Design:** Cross-sectional population-representative survey.

**Setting:** The National Health and Nutrition Examination Survey (NHANES, 2009-2012), a stratified multistage probability sample of the US population.

**Participants:** NHANES participants aged 18-69 years (n=8,237, representing 202,314,000 individuals).

**Exposures:** Demographic, socioeconomic, behavioral, anthropometric, metabolic, and clinical characteristics.

**Main outcomes:** Oral microbiome, characterized through 16S rRNA sequencing. Microbiome metrics were alpha diversity (number of observed Amplicon Sequence Variants [ASV], Faith’s Phylogenetic diversity, Shannon-Weiner Index, and Simpson Index); beta diversity (unweighted UniFrac, weighted UniFrac, and Bray-Curtis dissimilarity); and prevalence and relative abundance at taxonomic levels (phylum through genus). Analyses accounted for the NHANES complex sample design.

**Results:** Among US adults aged 18-69 years, the oral microbiome encompassed 37 bacterial phyla, 99 classes, 212 orders, 446 families, and 1,219 genera. Five phyla— *Firmicutes, Actinobacteria, Bacteroidetes, Proteobacteria,* and *Fusobacteria* and six genera—*Veillonella, Streptococcus, Prevotella7, Rothia, Actinomyces, and Gemella*, were present in nearly all US adults (weighted-prevalence >99%). These genera also were the most abundant, accounting for 65.7% of abundance. Observed ASVs showed a quadratic pattern with age (peak at 30 years), was similar by sex, significantly lower among non-Hispanic White individuals, and increased with higher body mass index (BMI) categories, alcohol use, and periodontal disease severity. All covariates together accounted for a modest proportion of oral microbiome variability, as measured by beta diversity (unweighted UniFrac=8.7%, weighted UniFrac=7.2%, and Bray-Curtis=6.3%). By contrast, relative abundance of a few genera explained a high percentage of variability in beta diversity (weighted UniFrac: *Aggregatibacter*=22.4%, *Lactococcus*=21.6%, *Haemophilus*=18.4%). Prevalence and relative abundance of numerous genera were significantly associated (Bonferroni-corrected Wald-p<0.0002) with age, race and ethnicity, smoking, BMI categories, alcohol use, and periodontal disease severity.

**Conclusions:** We provide a contemporary reference standard for the oral microbiome of the US adult population. Our results indicate that a few genera were universally present in US adults and a different set of genera explained a high percentage of oral microbiome diversity across the population.

## BACKGROUND

The community of bacteria in the human oral cavity, the oral microbiome, is increasingly recognized to play a role in health and disease.^1–3^ The oral microbiome is involved in digestion and metabolism of nutrients (e.g., dietary nitrates, carbohydrates) and carcinogens (e.g., conversion of ethanol to acetaldehyde), blood pressure and metabolic homeostasis,^1,4,5^ and regulation of oral and systemic immunity and inflammation.^1,4^ Importantly, certain oral bacteria contribute to periodontitis and poor oral health,^6^ and alterations in the oral microbiome have been associated with several chronic diseases, including cancer (upper-aerodigestive, colorectal), diabetes, cardiovascular disease, and neurological disorders.^1,3,4,7–11^

Understanding the relevance of the oral microbiome in health and disease requires a population reference standard of the composition of the oral bacterial ecosystem and prevalence, relative abundance, and interrelationships of taxa, established through characterization of the oral microbiome in a representative sample/population. Such a reference standard could serve as a critical comparator to understand the disease relevance of alterations in the oral microbiome. However, few population-representative characterizations exist in the literature.^12^ Most prior studies of the oral microbiome have been conducted in non-representative groups or highly selective subgroups. ^2,13–15^

Here, we address the lack of population-representative studies through characterization of the composition and diversity of the oral microbiome as well as demographic, socioeconomic, behavioral, metabolic, and clinical correlates of the oral microbiome in a representative sample of the US adult population aged 18-69 years.^16^

## METHODS

### Study population and biospecimens

We conducted this study using data from the National Health and Nutrition Examination Survey (NHANES), a complex stratified multistage cluster probability sample of the civilian, non-institutionalized US population.^17^ The survey includes a household interview component for collection of data on demographics, diet, tobacco use, and medical history and a mobile examination center (MEC) component for health, dental, anthropometric, and biochemical examinations and biospecimen collection.^17^

We conducted this study using oral rinse samples collected for oral human papillomavirus (HPV) evaluations in two consecutive NHANES cycles (2009-2010 and 2011-2012).^18^ Briefly, NHANES incorporated an oral HPV protocol in four cycles (2009-2016) for participants aged 14-69 years.^19,20^ Response rates for the MEC component were 68.5% and 69.5% in 2009-2010 and 2011-2012, respectively. The NHANES protocol was approved by the National Center for Health Statistics (NCHS) Ethics Review Board (ERB)^21^ and all participants provided written informed consent.

### Biospecimens and laboratory methods

Participants were provided a 10 mL oral rinse (Scope or saline) sample and completed 3 alternating swish and gargle cycles of 5 seconds each. Oral rinses were shipped weekly to the Gillison laboratory (Ohio State University) and stored at 4°C until further processing.^18^ Oral rinses were pelleted, lysed, and digested using DNase-free RNase A and proteinase K. DNA was extracted on the QIAsymphony SP instrument using the QIAGEN Virus/Bacteria Midi kit and Pathogen Complex 800 program and stored at -20°C (**Supplemental methods**).^18^

### 16S rRNA gene sequencing

Extracted DNA from n=9,997 participants was shipped to the Knight laboratory (University of California, San Diego) on dry ice and the V4 region of the 16S rRNA gene was PCR-amplified using the 515F-806R primers and sequenced using Illumina HiSeq 2500 with paired-end sequencing of 125 bp;^22^ however, only data from forward sequencing runs were used for analyses due to a lack of overlapping reads. A total of 19 individual HiSeq sequencing runs were conducted, with repeated sequencing of runs, plates, or plate-lanes, as necessary (**Supplemental methods**).

Of note, NCHS/NHANES policy at the time (circa 2015) did not allow targeting human DNA for sequencing due to participant confidentiality concerns, which precluded metagenomic sequencing. Consequently, our data lack species-level resolution and imputed bacterial functionality. Also, there are geographic bacterial niches within the oral cavity;^1,2,23,24^ thus, our results from oral rinses need to be interpreted as the generalized/sentinel microbiome profile of the biofilms within the oral cavity.

Details regarding quality control (QC) samples are described in the **Supplemental methods** QC analyses showed acceptable test-retest reproducibility; importantly, given no evidence for plate/batch effects we did not employ batch normalization procedures (**Figures S1-S2**).

### Bioinformatics methods and microbiome metrics

The FASTQ files were demultiplexed using QIIME 1.^25^ The forward reads from each sequencing run were processed using DADA2 and Amplicon Sequence Variants (ASV) were predicted.^26,27^ The phylogenetic tree was generated using QIIME 2 and taxonomy was assigned using the SILVA v123 database.^28,29^ For primary analyses, we created an ASV table that filtered out a non-bacterial ASV (SV1032, mapping to a mitochondrial pseudogene) that was identified in data quality checks (**Supplemental methods)**.

The resulting ASV tables were rarefied 10 times at 10,000 reads per sample and averaged for calculation of alpha and beta diversity metrics. We calculated four alpha diversity metrics using ASV tables: number of observed ASVs, a measure of richness of samples; Faith’s Phylogenetic Diversity [Faith’s PD], a phylogeny-weighted richness; Shannon-Weiner Index, a measure of evenness of samples; and Simpson Index, a measure of both richness and evenness of samples (**Supplemental Methods**).

We also calculated three beta diversity distance matrices for n x n comparisons across participants as measures of differences in the bacterial community: unweighted UniFrac, which considers presence/absence of bacterial ASVs to estimate phylogeny-weighted distance between participants; weighted UniFrac, which considers relative abundance of bacterial ASVs to estimate phylogeny-weighted distance between participants; and Bray-Curtis dissimilarity, which considers relative abundance of bacterial ASVs without phylogenetic weighting (**Supplemental Methods**).

Taxonomic analyses were based on non-rarefied data but were restricted to samples with at least 10,000 reads. Bacterial taxonomy was characterized across phylum (L2), class (L3), order (L4), family (L5), and genus (L6) levels, with data expressed as prevalence (presence/absence) and relative abundance (ratio of the number of each taxon-specific sequences and sum of all taxon-specific sequences in an individual). We primarily present results at the phylum (L2) and genus (L6) levels; characterizations at all levels are presented in **Tables S1-S6**.

### Statistical methods

All analyses accounted for the NHANES complex sampling design through the use of the cycle-specific NHANES sample weights (examination weights adjusted for non-response) and the strata and multistage clustered sample design. We created post-stratification adjustments to the cycle-specific examination weights to account for non-availability of oral microbiome results (18.4%) from MEC participants aged 18-69 years (**Table S7**). Specifically, NHANES MEC weights were post-stratified to the age by sex by race/ethnicity cycle-specific distribution of the US population. Post-stratified weights were divided by two given the use of two NHANES cycles.

We describe the structure and diversity of the oral microbiome through descriptive statistics. Results are presented as percentages, means, medians, and quantiles, as appropriate. Taxa by taxa relative abundance correlations were calculated using sample weighted Pearson’s correlation coefficients (**Tables S8-S12**). Population totals were estimated using post-stratified weights.

Analyses considered demographic and socioeconomic factors (age, modeled using restricted 5-knot cubic regression splines, sex, race and ethnicity, education, marital status, income-to-poverty ratio), anthropometrics (measured body mass index [BMI], categorized), risk behaviors (smoking, alcohol use), medical conditions (diabetes and hypertension), oral health (periodontal disease, tooth count, and edentulism), and use of prescription medications within the past 30 days (antibiotics, antilipidemics, respiratory inhalants, and for gastroesophageal reflux). Race and ethnicity was self-reported as Mexican American, Other Hispanic, non-Hispanic White, non-Hispanic Black, and non-Hispanic other race and ethnicity (including Native American, Pacific Islander, Asian and multi-racial). These variables were chosen a priori for adjustment in statistical models (variable details and code in **Table S13**).

Correlates of the four alpha diversity metrics were evaluated using multiple linear regression with each metric as the dependent variable. Results from these models are expressed as predictive margins, analogous to adjusted means.^30^ For the three beta diversity matrices, we estimated sample weighted principal coordinate analysis (PCoA) vectors and utilized the first 100 PCoAs (scree plots in **Figures S3**) as outcomes in linear regression models.

We utilized *FastAdonis*^31^ to estimate partial and full-model explained variability in the beta diversity matrices from each variable and combinations of variables using the ANOVA principle, while accounting for the complex design. Similarly, we estimated the contribution of each genus to variability in the beta diversity matrices, unadjusted for covariates or correlations across genera.^31^

Our analyses of covariate associations examined genus-level associations with prevalence of genera using binary logistic regression, results presented as odds ratio [OR]. Associations with relative abundance used Poisson regression, with genus-level sequence counts as the outcomes and the total sequence counts as the offset; results are presented as risk ratios [RR]. We note that microbiome relative abundance data seldom follow a Poisson distribution and show zero inflation, which results in variance inflation. It is, however, difficult to implement zero-inflated regression (e.g., zero-inflated negative binomial or Poisson regression) while accounting for the NHANES design because of recognized issues with model convergence. Thus, we utilized complex survey design-based Poisson regression, which provides robust variances that account for extra-Poisson variability. Logistic and Poisson regression models were adjusted for covariates noted herein, except respiratory inhalant drugs (excluded due to sparse sample sizes).

Associations with alpha and beta diversity utilized two-sided Wald-p<0.05 for statistical significance. Analyses of the genus-level prevalence and relative abundance were restricted to genera with at least 1% prevalence (n=229 genera) and utilized a Bonferroni-corrected Wald-p-value threshold of 0.0002 (=0.05/229) for statistical significance. Analyses were conducted in SAS-callable SUDAAN version 10.4.1 (Raleigh, North Carolina) and R v4.2.0.

## RESULTS

Among 8,237 US adults aged 18-69 years (representing 202,314,000 individuals), the oral microbiome encompassed 37 bacterial phyla, 99 classes, 212 orders, 446 families, and 1,219 genera. Five phyla—*Firmicutes, Actinobacteria, Bacteroidetes, Proteobacteria,* and *Fusobacteria* **(Figure 1A**) and six genera— *Veillonella, Streptococcus, Prevotella7, Rothia, Actinomyces, and Gemella*, (**Figure 1B**) were present in nearly all US adults (weighted prevalence >99%). These genera also had the highest relative abundances (**Figure 1B inset**), collectively accounting for 65.7% of the genus-level total abundance—*Streptococcus* (32.6%), *Rothia* (11.5%), *Prevotella7* (8.0%), *Veillonella* (6.3%), *Gemella* (4.8%), and *Actinomyces* (2.7%). At the population-level, however, genus-level prevalence was only moderately correlated with its relative abundance (rho=0.35), as most prevalent genera had very low relative abundances.

**Figure 1:**
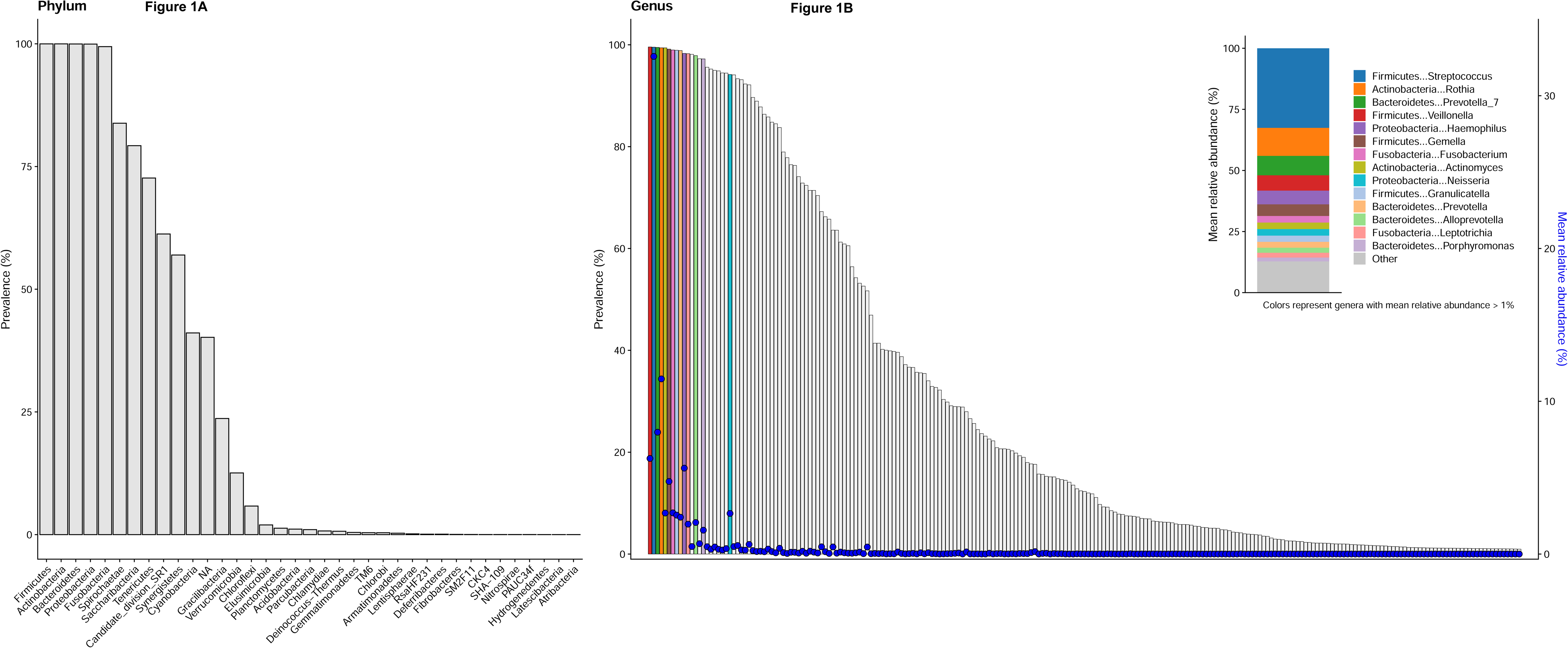
Shows the prevalence of phyla (**panel A**) and prevalence of genera (**panel B**) in the oral microbiome among US adults aged 18-69 years, NHANES 2009-2012; bars represent weighted percent prevalence and circles represent weighted mean relative abundance. The inset in panel B shows the proportionate weighted mean relative abundance for genera with >1% relative abundance and for the remaining genera.

### Alpha diversity associations

The estimated mean oral microbiome alpha diversity in US adults aged 18-69 years was 128 (95% CI=126-130) for observed ASVs, 14.5 (95% CI=14.3-14.7) for Faith’s PD, 4.58 (95% CI=4.55-6.61) for the Shannon-Weiner Index, and 0.90 (95% CI=0.89-0.90) for the Simpson Index.

Across years of age, the number of observed ASVs showed a non-linear quadratic pattern, with an increase from ages 18 to 30 years, peak at age 30 years, and subsequent decline after 30 (**Figure 2**). The number of observed ASVs was similar for men and women. Compared to non-Hispanic White individuals (124 ASVs), observed ASVs were significantly higher for individuals of all other race and ethnicity groups (**Figure 3**, Mexican American (137), other Hispanic (132), non-Hispanic Black (134), and other non-Hispanic (131), including non-Hispanic Asian (127), a group first oversampled in NHANES 2011-2012). Observed ASVs decreased with increasing education and increased with increasing BMI, alcohol use (among ever drinkers), and increasing severity of periodontal disease (**Figure 3**). Notably, the number of observed ASVs was substantially lower in edentulous individuals (75 vs. population average=128). Use of prescription medications over the past 30 days, including antibiotics, antilipidemic medications, and medications for hypertension and gastroesophageal reflux were associated with lower observed ASVs, while inhaled respiratory drug use did not alter this metric (**Figure 3**). Similar associations were observed for the other alpha diversity metrics (**Figures S4-S6**).

**Figure 2:**
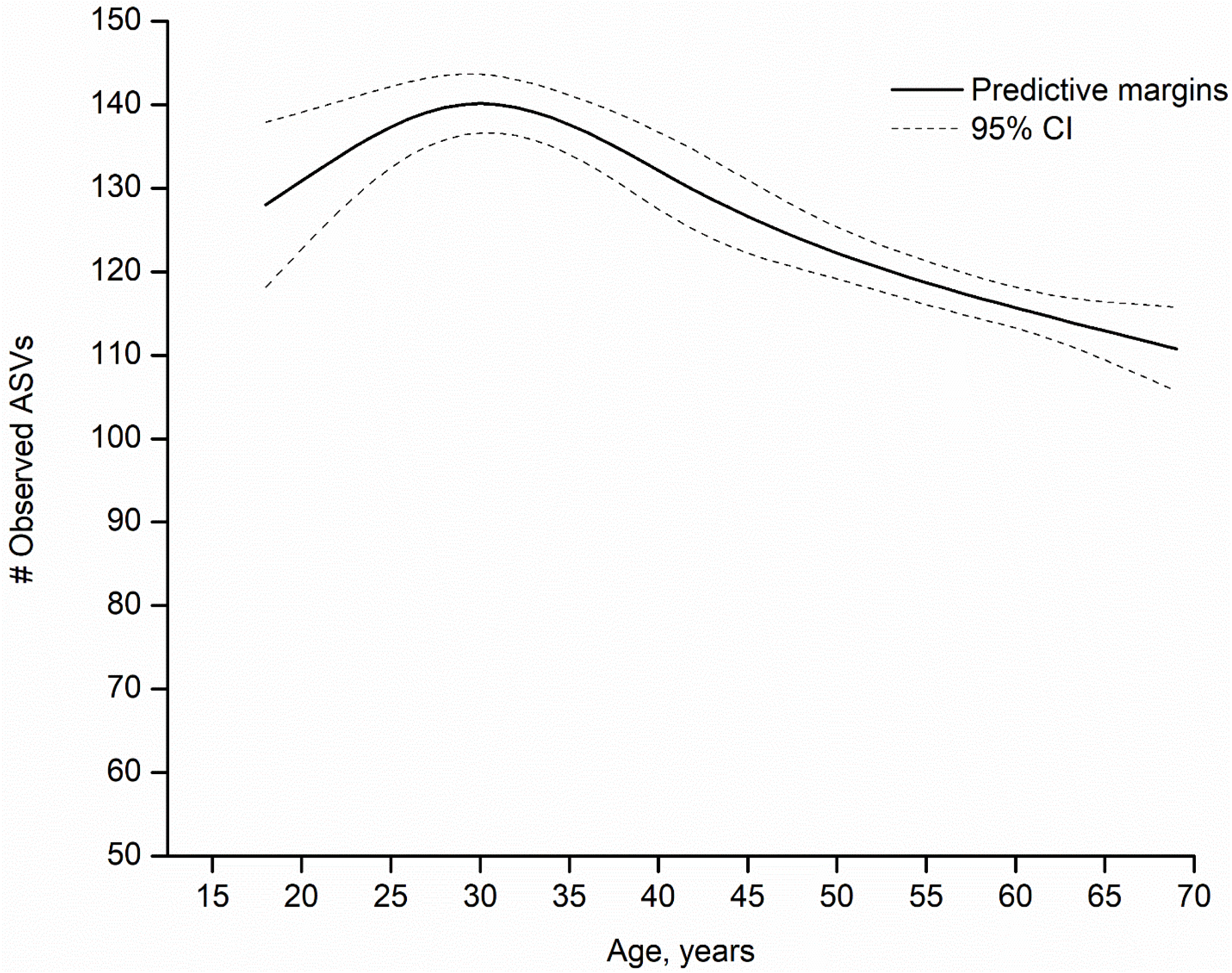
Shows the adjusted predictive margins (solid line) and 95% confidence intervals (dashed lines) for the observed number of amplicon sequence variants (ASVs) by age (in years) in the oral microbiome of US adults aged 18-69 years, NHANES 2009-2012. Age was modeled through 5-knot restricted cubic splines. Predictive margins were estimated in survey-design adjusted linear regression models, with concomitant adjustment for sex, race and ethnicity, education, marital status, income-to-poverty ratio, measured body mass index [BMI] categories, risk behaviors (smoking, alcohol use), medical conditions (diabetes and hypertension), oral health (periodontal disease, tooth count, and edentulism), and use of prescription medications within the past 30 days (antibiotics, antilipidemics, respiratory inhalants, and for gastroesophageal reflux). Race and ethnicity was self-reported and used as categorized by NHANES. Please see Table S13 for variable definitions/code.

**Figure 3:**
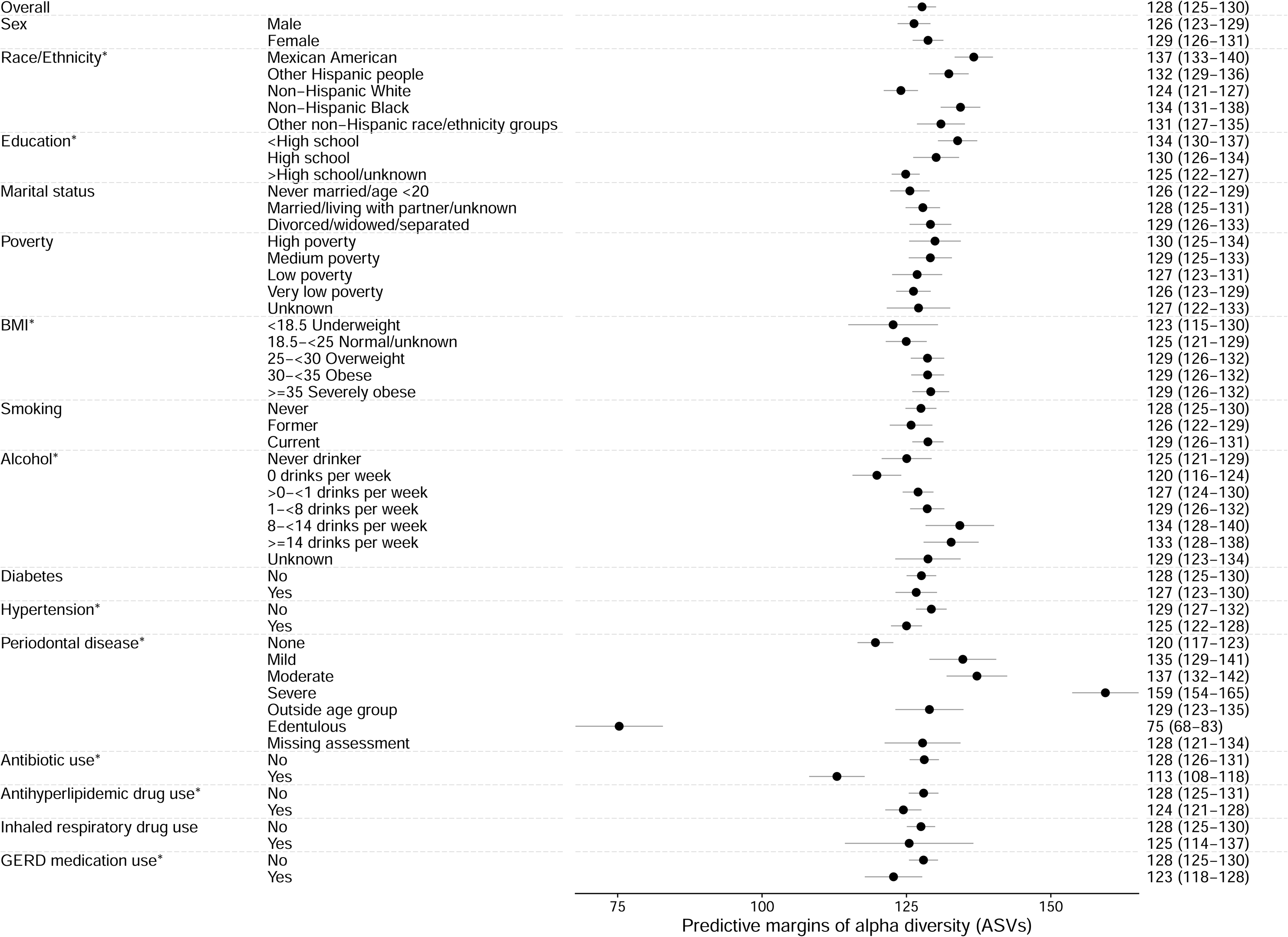
Shows the adjusted predictive margins (circles) and 95% confidence intervals (error bars) for the observed number of amplicon sequence variants (ASVs) in the oral microbiome of US adults aged 18-69 years (NHANES 2009-2012) by each covariate. Predictive margins were estimated in survey-design adjusted linear regression models, with concomitant adjustment for age (modeled as 5-knot restricted cubic splines), sex, race and ethnicity, education, marital status, income-to-poverty ratio, measured body mass index [BMI] categories, risk behaviors (smoking, alcohol use), medical conditions (diabetes and hypertension), oral health (periodontal disease, tooth count, and edentulism), and use of prescription medications within the past 30 days (antibiotics, antilipidemics, respiratory inhalants, and for gastroesophageal reflux). Race and ethnicity was self-reported and used as categorized by NHANES. Please see Table S13 for variable definitions/code. An asterisk denotes p<0.05 for the covariate in the adjusted model.

### Beta diversity associations

A modest percentage (<9%) of the overall variability in beta diversity was explained by the demographic, socioeconomic, behavioral, and health covariates— 8.7%, 7.2%, and 6.3%, for unweighted UniFrac, weighted UniFrac, and Bray-Curtis matrices, respectively (**Figure 4A-B**). Dominant factors in weighted UniFrac analyses for beta diversity variability were periodontal disease (3.3%), age (1.1%), and smoking (2.4%). These covariates and antibiotic use were also significantly associated with the first PCoA axis across the three beta diversity matrices (**Tables S14-S16**).

**Figure 4:**
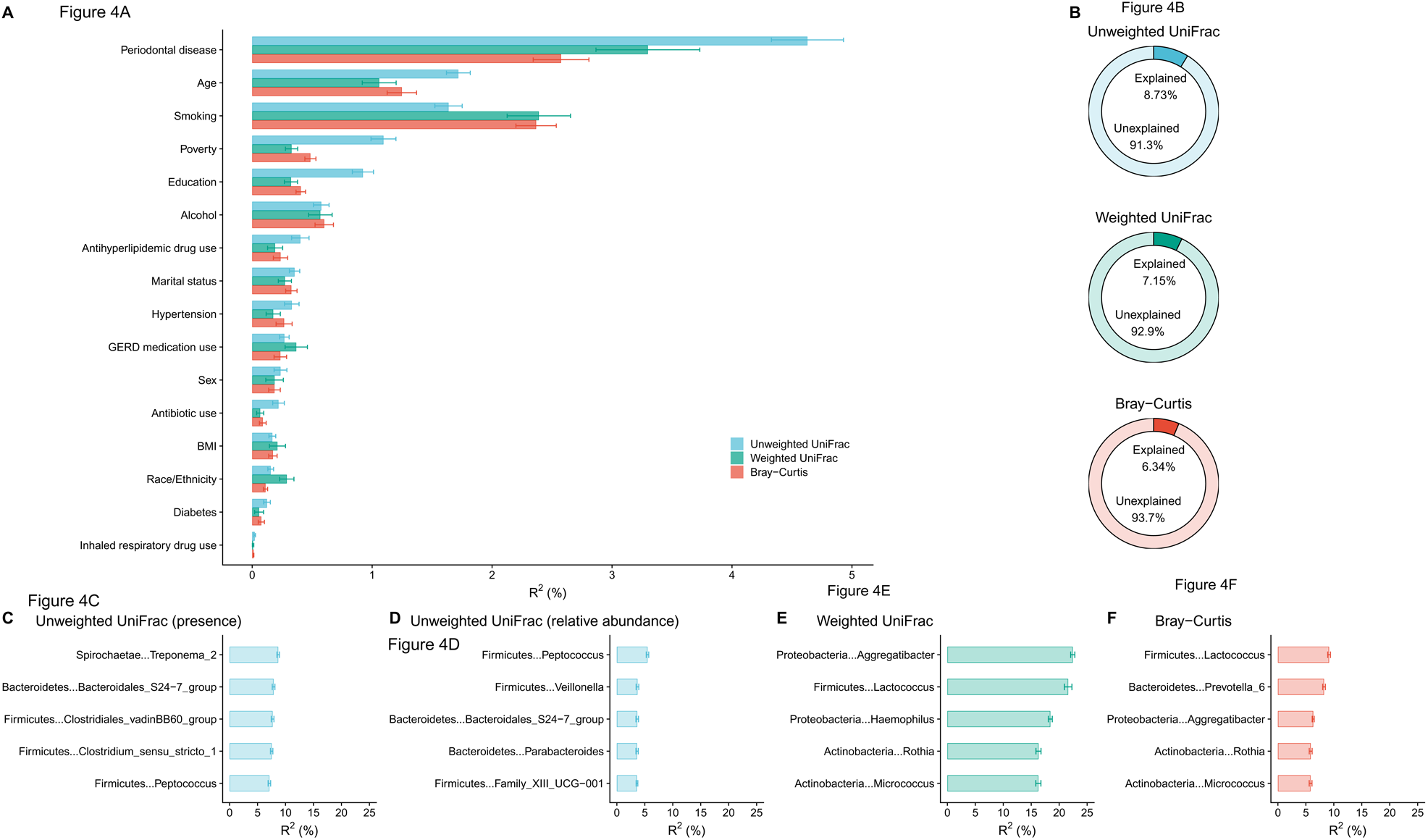
**Panel A** shows the covariate-specific (horizontal bars) and overall (horizontal bars) proportion of variability in beta diversity matrices (unweighted UniFrac, weighted UniFrac, and Bray-Curtis) in the oral microbiome of US adults aged 18-69 years, NHANES 2009-2012. The fastAdonis algorithm was used for estimation. The overall estimate was based on models that incorporated concomitant adjustment for age (modeled as 5-knot restricted cubic splines), sex, race and ethnicity, education, marital status, income-to-poverty ratio, measured body mass index [BMI] categories, risk behaviors (smoking, alcohol use), medical conditions (diabetes and hypertension), oral health (periodontal disease, tooth count, and edentulism), and use of prescription medications within the past 30 days (antibiotics, antilipidemics, respiratory inhalants, and for gastroesophageal reflux). Race and ethnicity was self-reported and used as collected by NHANES. Covariate-specific estimates are unadjusted. Please see Table S13 for variable definitions/code. The proportion of variability in the unweighted UniFrac beta diversity matrix from prevalence of each genus is shown in **panel C** and from relative abundance of key genera is shown in **panel D**. **Panel E** shows the proportion of variability in the weighted UniFrac beta diversity matrix from relative abundance of key genera. **Panel F** shows the proportion of variability in the Bray-Curtis beta diversity matrix from relative abundance of key genera. The fastAdonis algorithm was used for estimation and genus-specific estimates are unadjusted for covariates or correlations across prevalence or relative abundance of genera.

By contrast, a few genera explained a surprisingly high percentage of variability in weighted UniFrac in particular—relative abundance of 3 genera: *Aggregatibacter* (22.4%), *Lactococcus* (21.6%), and *Haemophilus* (18.4%) (**Figure C-F)**. Importantly, these genera had marked variation in average prevalence and relative abundance compared to each other—*Aggregatibacter* prevalence=67.3%, relative abundance=0.5%; *Lactococcus* prevalence=5.5%, relative abundance=0.007%; and *Haemophilus* prevalence=98.3%, relative abundance=5.6%—underscoring their influence on beta diversity, particularly weighted UniFrac, regardless of prevalence or relative abundance.

### Genus-level associations

The prevalence and/or relative abundance of genera with major contributions to variability in weighted UniFrac (*Aggregatibacter, Lactococcus, Haemophilus)* also had associations with several covariates. For example, prevalence of *Aggregatibacter* was significantly associated with race and ethnicity, smoking, periodontal disease, and antibiotic use; relative abundance of *Haemophilus* was 5-times higher among antibiotic users than non-users and 3-times higher among obese individuals compared to normal weight individuals (**Tables S17-S18).**

Numerous bacterial genera were significantly associated with particular covariates, some with multiple covariates, at a Bonferroni-corrected p-value threshold (p<0.0002) (**Figure 5A**). For example, prevalence of *Alloscardovia* and of *Lachnospiraceae* was significantly associated with age, race and ethnicity, smoking, alcohol use, and periodontal disease. Likewise, relative abundance of *Castellaniella* and *Phascolarctobacterium* was significantly associated with age, race and ethnicity, periodontal disease, alcohol use, and BMI. The genera associated with each covariate generally differed for prevalence and relative abundance, except for periodontal disease where both measures were associated with mostly the same genera (**Figure S7**).

**Figure 5:**
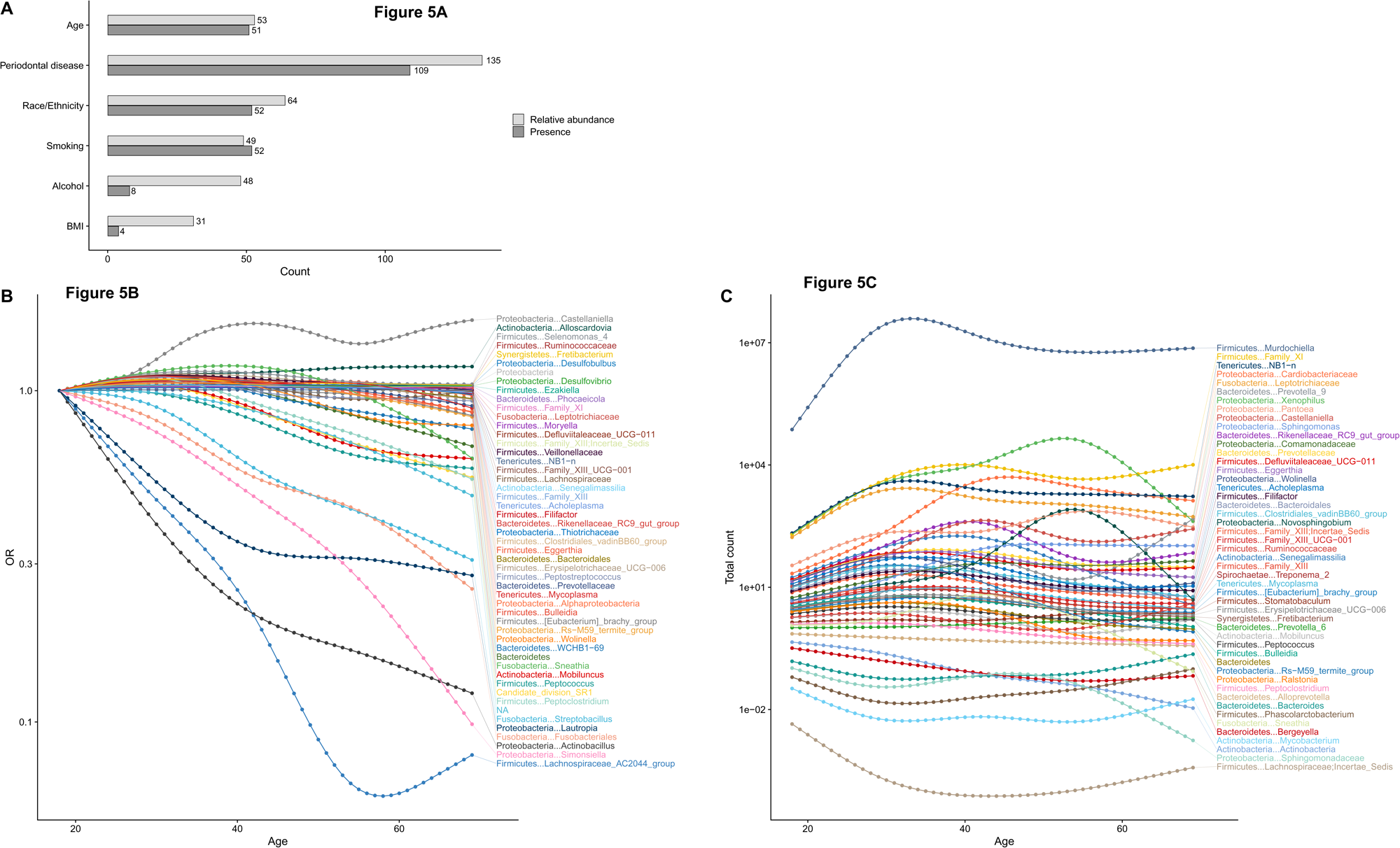
Panel **A** shows for key covariates the number of genera with statistically significant (at a Bonferroni-corrected p<0.0002) associations with genus-specific relative abundance (light grey bars) and genus-specific prevalence (dark grey bars) for ages 18-69 years, NHANES 2009-2012. Associations with relative abundance were estimated in Poisson regression models and with prevalence were estimated in binary logistic regression models. **Panel B** shows the adjusted odds ratio (OR) by age (18-69 years) for prevalence of genera with statistically significant associations with age at a Bonferroni-corrected p<0.0002. **Panel C** shows the model-predicted genus-specific sequence counts by age (18-69 years) for genera with statistically significant association of genus-specific relative abundance with age at Bonferroni-corrected p<0.002. All models included concomitant adjustment for age (modeled as 5-knot restricted cubic splines), sex, race and ethnicity, education, marital status, income-to-poverty ratio, measured body mass index [BMI] categories, risk behaviors (smoking, alcohol use), medical conditions (diabetes and hypertension), oral health (periodontal disease, tooth count, and edentulism), and use of prescription medications within the past 30 days (antibiotics, antilipidemics, and for gastroesophageal reflux). Race and ethnicity was self-reported and used as categorized by NHANES. Please see Table S13 for variable definitions/code.

Similar to alpha diversity, prevalence and relative abundance of several genera were non-linearly associated with age, with varying age peaks (**Figures 5B and 5C**). Prevalence of *Desulfomicrobium* was >5-times higher in each of the four race and ethnicity groups compared to non-Hispanic White individuals (**Table S17**). Prevalence of *Metascardovia* was 13-times higher in current smokers vs. never smokers. Current smoking was also associated with increased prevalence of several genera in the *Bifidobacteriaceae* family (*Metascardovia, Scardovia, Parascardovia, Aeriscardovia, Alloscardovia*). In contrast, prevalence of *Neisseria* was 3-times higher in never smokers than current smokers. Prevalence of *Castellaniella* increased with increasing alcohol use and was ∼5-times higher among heavy vs. never drinkers.

Strong associations were also observed for relative abundance (**Table S18**) by race and ethnicity (*Murdochiella* 5-18-times higher abundance in non-White races and ethnicities vs. non-Hispanic White individuals), smoking (*Metascardovia* 14-times higher abundance in current vs. never smokers), and BMI categories (*Coprobacter* 10-times higher abundance among adults with obesity vs. normal weight individuals).

Increasing severity of periodontal disease was generally associated with increased prevalence and increased relative abundance of several genera. Despite the lack of resolution in our results at the species level, both prevalence and relative abundance of red complex bacterial genera (*Porphyromonas, Tannerella,* and *Treponema*) were significantly higher among participants with severe periodontitis when compared to those with no periodontal disease (**Tables S17-S18**).

## DISCUSSION

Our study provides a contemporary reference standard for the oral microbiome of the US adult population aged 18-69 years during 2009-2012 (representing 202,314,000 individuals). We show that the oral microbiome is a complex ecosystem encompassing 37 phyla and over 1,000 genera. Yet, a limited set of genera (*Veillonella, Streptococcus, Prevotella7, Rothia, Actinomyces, and Gemella)* were observed in all adults (>99%) and accounted for a majority of the genus-level relative abundance (66%), indicating the existence of a limited universal oral microbiome in US adults.^2,12,13^ A different, limited set of genera (*Aggregatibacter, Lactococcus, Haemophilus*) were associated with a high proportion of variability in one measure of oral microbiome beta diversity (>60% in weighted UniFrac), indicating the existence of key genera that potentially influence oral microbiome diversity across individuals. A wide range of demographic, behavioral, and metabolic/clinical covariates (age, race and ethnicity, smoking, alcohol consumption, and periodontal disease) were consistently associated with multiple oral microbiome metrics—alpha diversity, beta diversity, and genus-level prevalence and relative abundance—underscoring potential modifiability of the oral microbiome.

Some of the alpha diversity, beta diversity, and genus-level associations we report have previously been noted in the literature and may have plausible biological explanations. ^32,33^ Such observations include enrichment of *Actinobacteria* among current smokers (particularly several genera in the *Bifidobacteriaceae* family)^32^ and red complex bacteria in those with severe periodontal disease.^33^ Nonetheless, several of our key observations remain unexplained. The non-linear associations of age with oral microbiome alpha diversity (peaking at age 30) and prevalence and relative abundance of several genera (varying age peaks), has not been previously reported. The numerous associations of race and ethnicity with alpha and beta diversity metrics and genus-level prevalence and relative abundance, although previously reported,^11,2^ were not explained by the socioeconomic, behavioral, anthropometric, metabolic, and clinical factors we included for statistical adjustment.^14,34^ Additional hypotheses and research are needed to identify potential biological or alternative explanations. For example, the non-linear association with age could arise from biological ageing or reflect birth cohort effects. However, we could not evaluate these birth cohort effects because the narrow calendar years in our study makes age equivalent to birth cohort. Likewise, associations with race and ethnicity could reflect multifactorial social determinants of health, such as early life exposures or access to health services,^34^ which were not considered herein.

Our observations suggest that the interpretation of alpha diversity as an indicator of health/disease may be context-specific. Although high alpha diversity (particularly of the gut microbiome) is generally believed to be an indicator of good health/outcomes,^35^ we show that oral microbiome alpha diversity (as measured by observed number of ASVs and after adjustment for a wide range of covariates) was higher among those with severe periodontal disease and overweight and obesity but substantially lower among edentulous individuals. The lower richness in edentulous individuals suggests the important role of teeth in creating diverse niches in the oral environment.

We note key study limitations and strengths. Our evaluation was restricted to 16S rRNA gene sequencing because metagenomic sequencing was precluded by NCHS policy. Thus, the data resolution is through the genus-level and we lacked species differentiation and imputed bacterial functionality, which metagenomic sequencing would have permitted. The strengths of our study are the population representative nature of the data, large sample size, and the comprehensive array of demographic, socioeconomic, and behavioral data and uniform, high-quality data, and unindicated (i.e., assessment independent of health status) health examinations.

In summary, we present the oral microbiome profile of US adults—colloquially, the mouth of America. Importantly, NCHS, CDC has made available all the results to the research community (publicly available data can be accessed at the NHANES website: https://wwwn.cdc.gov/Nchs/Nhanes/omp/ and data considered restricted can be accessed at NCHS Research Data Centers: https://www.cdc.gov/rdc/index.htm). Our comprehensive characterization herein, and the anticipated use of the data by the scientific community, portends deeper understanding of the role of the oral microbiome in health and disease.

## Supporting information

Supplement Index

## Data Availability

Unlinked public-use National Health and Nutrition Examination Survey (NHANES) data files are available on the NHANES website (https://www.cdc.gov/nchs/nhanes). The linked mortality (National Death Index) files are available in a public-use version, available online (https://www.cdc.gov/nchs/data-linkage/mortality.htm), and a restricted-use file available for analysis at the National Center for Health Statistics Research Data Center (https://www.cdc.gov/rdc/index.htm). The restricted-use mortality file was used for this analysis.

