## Supplement Index for "The mouth of America: the oral microbiome profile of the US population"

**Data supplement index for: “The mouth of America: the oral microbiome profile**

**of the US population.”**

**Item**

1. Supplemental methods (DNA extraction, sequencing, bioinformatics, and quality control)

2. Figures S1 and S2: relative abundance of major phyla and genera in quality control samples across sequencing plates/batches

3. Figure S3: Scree plot of principal coordinate analyses (PCoAs) for beta diversity matrices (Bray-Curtis, unweighted UniFrac, and weighted UniFrac)

4. Figures S4-S6: Results of associations of covariates with alpha diversity metrics (Faith’s PD, Shannon-Wiener Index, and Simpson Index)

5. Figure S7: Heatmap of associations of covariates with prevalence and relative abundance of genera

6. Table S1: Taxonomy key

7. Tables S2-S6: Prevalence at phylum (L2), class (L3), order (L4), family (L5), and genus (L6) levels

8. Table S7: Characteristics of NHANES 2009-2012 participants aged 18-69 years with or without oral microbiome data

9. Tables S8-S11Taxa x taxa correlations of relative abundance at phylum (L2), class (L3), order (L4), and family (L5)

10. Table S12: S11Taxa x taxa correlations of relative abundance at gen(L6) level

11. Table S13: Code and variable	definitions used in the analyses

12. Tables S14: Associations of covariates with first 100 principal coordinates for beta diversity matrices (Bray-Curtis) , unweighted UniFrac, and weighted UniFrac)

13. Tables S15: Associations of covariates with first 100 principal coordinates for beta diversity matrices (unweighted UniFrac)

14. Tables S16: Associations of covariates with first 100 principal coordinates for beta diversity matrices (weighted UniFrac)

15. Table S17: Associations of covariates with prevalence of genera, logistic regression

16. Table S18: Associations of covariates with relative abundance of genera, Poisson regression
