## Supplemental methods for "The mouth of America: the oral microbiome profile of the US population"

### National Health and Nutrition Examination Survey
### 2009-2010 and 2011-2012
### Oral Microbiome Data Documentation

First Published: October 2022 Last Revised: N/A

#### Component Description

It is increasingly recognized that alterations in the oral microbiome could be relevant for health and disease. Alterations in the oral microbiome are believed to have both local as well as systemic health effects. Characterization of the oral microbial profile in a representative sample of the US population, such as the National Health and Nutrition Examination Survey (NHANES), will help to better understand the association of perturbations in the oral microbiome. The rich data on demographics, behaviors, laboratory assessments, and health conditions in NHANES allows a comprehensive characterization of the oral microbiome in health and disease in the US population. Such a characterization could provide a population reference of the oral microbiome for future evaluations of associations with disease in the United States.

Oral microbiome testing was performed using oral rinse samples collected from NHANES participants in two consecutive NHANES cycles, 2009-2010 and 2011-2012. Results from the oral microbiome testing encompass classification of microbial sequences into amplicon sequence variants (ASVs) which were used to generate alpha-diversity metrics, beta-diversity matrices, and relative abundance/read count tables for each taxonomic level from phylum to genus.

This documentation contains relevant information for NHANES' oral microbiome datasets in 2009-2010 and 2011-2012. This general documentation describes the samples, laboratory methods, and bioinformatics procedures for these oral microbiome NHANES data files.

#### Eligible Samples

All examined participants aged 14-69 years in the 2009-2010 and 2011-2012 samples were eligible.

#### Description of Laboratory Methodology

DNA was extracted from the oral rinse samples using methods described in detail previously (https://wwwn.cdc.gov/nchs/data/nhanes/2009-2010/manuals/HPV.pdf)[1]. Seventy-two 96-well plates from 2009-2010 and sixty 96-well plates from 2011-2012 were created with extracted DNA from NHANES oral rinse samples, blank samples, and two types of quality control samples (oral artificial community and artificial gut samples).

These plates were shipped on dry ice to the Knight laboratory at the University of California, San Diego, CA. Upon receipt, the plates were stored at -20°C prior to processing. Polymerase chain reaction (PCR) amplification and sequencing were performed as published previously[2] and as detailed on the Earth Microbiome website (https://earthmicrobiome.org/protocols-and-standards/16s/). Specific details are below:

1. Using an EpMotion (Eppendorf) liquid handler, the PCR reaction mixture was created using the following reagents and volumes. Each sample was amplified in 3 replicate 25-µL PCR reactions.

| Reagent | Volume |
|---|---|
| PCR-grade water from Sigma (cat. No. W3500) or MoBio (cat. No. 17000-11) | 13.0 µL |
| Platinum Hot Start PCR master mix (2x) from Thermofisher (cat. no. 1300014) | 10.0 µL |
| Forward primer (10 µM) – 515F[3] | 0.5 µL |
| Reverse primer (10 µM) – 806R[4] | 0.5 µL |
| Template DNA | 1.0 µL |
| **Total reaction volume** | **25.0 µL** |

2. The thermocycler had the following conditions for the 16S V4 515F-806R primers with ~390 bp amplicons.

| Temperature | Time, 384-well | Repeat |
|---|---|---|
| 94°C | 3 min | |
| 94°C | 60 sec | 35 times |
| 50°C | 60 sec | 35 times |
| 72°C | 105 sec | 35 times |
| 72°C | 10 min | |
| 4°C | Hold | |

3. The triplicate PCR reactions for each sample were pooled into a single volume (75 µL).

4. The amplicons from each sample were run on an agarose gel.

5. The amplicons were quantified using the Quant-iT PicoGreen dsDNA Assay Kit (ThermoFisher/Invitrogen cat. no. P11496) following the manufacturer's instructions.

6. An equal amount of amplicon from each sample (240 ng) were combined into a single, sterile tube.

7. The amplicon pool was cleaned using the MoBio UltraClean PCR Clean-Up Kit (cat. no. 12500) following the manufacturer's instructions.

8. The concentration and A260/A280 ratio of the final pool was measured.

9. An aliquot was sequenced using the Illumina HiSeq 2500 (2x125 bp) following the manufacter's instructions.

Nineteen individual HiSeq runs were used. Any HiSeq run or lane that failed (i.e., no or < 100 reads in all samples), in addition to participant failed samples within runs, were rerun on a subsequent HiSeq run. Data for quality control samples are available in separate datasets. If samples from the same participant were included in two different runs, only samples in the final (redo) run were maintained. The mapping file was provided as the input for all different processing methods.

#### Bioinformatics Procedures

The following sections refers to all bioinformatics procedures and includes code. Please refer to the flow chart for an illustration of the workflow.

##### 1. Demultiplexed raw sequence files

The raw sequence files for each run includes three multiplexed files (forward read file, reverse read file, and barcode file). The demultiplexing process (i.e., the process of associating 16S rRNA gene sequences with the sample that they were derived from based on DNA barcodes) was performed in QIIME1[5] using the command below for each run, which generates separate forward and reverse read files for each individual. This process generated a total of 10,442 forward fastq files and 10,440 reverse fastq files.

QIIME1 Command logs:

```
split_libraries_fastq.py –i $forward_fastq –m $mapping –o $forward_folder –b $barcode –barcode_type 12 –
rev_comp_barcode –rev_comp_mapping_barcodes –store_demultiplexed_fastq
split_sequence_file_on_sample_ids.py –i $forward_folder/seqs.fastq –o $forward_folder/out/ --file_type fastq
for f in *.fastq; do mv $f ${f/.fastq/_R1.fastq}; done

split_libraries_fastq.py –i $reverse_fastq –m $mapping –o $reverse_folder –b $barcode –barcode_type 12 –
rev_comp_barcode –rev_comp_mapping_barcodes – store_demultiplexed_fastq
split_sequence_file_on_sample_ids.py –i $reverse_folder/seqs.fastq –o $reverse_folder/out/ --file_type fastq
for f in *.fastq; do mv $f ${f/.fastq/_R2.fastq}; done
```

##### 2. Pipelines

The reads from the raw sequence files (i.e., the sequences generated from all individuals) were processed using the DADA2 pipeline [version 1.2.1])[6]. Since the generated reads were only 125 bp, there was no overlap between the forward and reverse reads to merge paired ends. Therefore, we only used the forward reads for analysis. Each sequencing run (i.e., each of the 19 individual HiSeq runs) was processed independently and then results were merged. The DADA2 pipeline predicts ASVs after modeling and correcting sequencing errors of microbiome data. Taxonomy was assigned by using SILVA v123 database[7].

##### 3. Phylogenetic trees

A phylogenetic tree is a representation of the hypothesized evolutionary relationships between ASVs or more generally, any biological entities. For DADA2, to create the phylogenetic tree, first, the sequence file, which includes all ASVs, was processed by DADA2. Then the phylogenetic tree, including all the ASVs, was generated by running the QIIME2[8] command.

QIIME2 Command logs:

```
    mafft –preservecase –thread -4 result.fasta > result-aligned.fasta
    qiime tools import –input-path result-aligned.fasta –output-path aligned-sequences.qza –type
FeatureData[AlignedSequence]
```

qiime alignment mask --i-alignment aligned-sequences.qza --o-masked-alignment masked-aligned-sequences.qza
qiime phylogeny fasttree –i-alignment masked-aligned-sequences.qza –o-tree unrooted-tree.qza
qiime phylogeny midpoint-root –i-tree unrooted-tree.qza –o-rooted-tree rooted-tree.qza
qiime tools export rooted-tree.qza –output-dir ./

#### 4. Amplicon sequence variant (ASV) feature table

The ASV feature table is a table that presents the read counts for each ASV for each individual. Typically, the ASV feature tables are stored in the BIOM[9] format. For sequence-based pipelines (DADA2), each ASV corresponds to one exact 16S rRNA gene sequence.

For the DADA2 pipeline, the ASV feature table was generated from DADA2 (1.2.1). Taxonomy assignment was based on SILVA v123 database. The ASV feature table includes 10,442 samples and 41,378 ASVs.

After analyzing the DADA2 data, it was noted that two distinct clusters in the beta diversity data were observed. Upon further analysis, one ASV was identified to be related to the clustering (SV1032). SV1032 is a non-bacterial taxon with the following sequence:

TGAACCCAAGCTAATAGAGACTGGCGTAAAGAATGTTTTACATTATCCCTCAATAAAGCTA
AATTTCACCTAAGTTGTAGAAAACCCTAGTTGATATAAAACAAACTACGAAAGTGGCTT

We therefore created two ASV feature tables to remove this clustering: 1) ASV feature table without SV1032 (RSV); and 2) ASV feature table removing all taxa without a phylum level identification (RB). For RB, we removed 2750 OTUs with the taxonomic assignment "Bacteria; NA; NA; NA; NA; NA." For RSV and RB, the specified sequences were removed from the ASV tables and the BIOM table was regenerated. These two feature tables were processed in parallel and all outputs described below are available for each feature table.

#### 5. Alpha diversity

Alpha diversity data and variable lists are available as public use files.

The alpha diversity is a measurement of diversity of the microbiome within a single sample (i.e., within a participant), typically representing community richness and/or community evenness. Alpha diversity is commonly measured with metrics including observed ASVs, Faith's Phylogenetic Diversity[10], the Shannon-Weiner index[11], and the Simpson Index[12]. These four alpha diversity metrics were calculated based on rarefaction values (i.e., random even subsampling of counts in a feature table without replacement to a specified total frequency) from 2,000 to 10,000. Faith's Phylogenetic Diversity utilizes phylogenetic tree information while the observed ASVs, Shannon-Weiner and Simpson indices do not use phylogenetic tree information. The observed ASVs and Faith's Phylogenetic Diversity metric measures richness only. The Shannon-Weiner and Simpson indices measures richness and evenness.

All alpha diversities for each pipeline were summarized in one table using an internal R script. In each alpha diversity table, each row represents one human sample and each column represents one alpha diversity value for the given sample. For example, D2_ObservedOTUs_2000_0 means DADA2 pipeline for Observed ASV counts with rarefaction at 2000 reads/sample for the first resampling (repetition = 0).

QIIME1 Command Logs:

```
multiple_rarefactions.py –i dada2_mod_final.biom –m 2000 –x 10000 –s 2000 –o alphaout
alpha_diversity.py –i alphaout/ -o alphaout/alpha_div –t dada2.tre –m
PD_whole_tree,observed_species,shannon,simpson
collate_alpha.py –i alphaout/alpha_div/ -o alphaout/alpha_collated/
```

#### 6. Beta diversity

Beta diversity data are available as public use files.

The beta diversity is a measurement of the microbiome diversity between samples (i.e., participants), represented in these datasets by pairwise dissimilarity of samples. Beta diversity is measured between all pairs of samples with metrics including unweighted UniFrac, weighted UniFrac[13], and Bray-Curtis dissimilarity[14]. Measurement of beta diversity between all pairs of samples is represented in a distance matrix.

The ASV feature tables were rarefied at 10,000 reads/sample by using QIIME command beta_diversity. For unweighted UniFrac distance and weighted UniFrac distance, phylogenetic trees were used. The Bray-Curtis dissimilarity index does not use a phylogenic tree. Similar procedures were applied to all three pipelines.

QIIME1 Command logs:

```
single_rarefaction.py –i dada2_mod_final.biom –o dada2_mod_final_even10k.biom –d 10000
beta_diversity.py –i dada2_mod_final_even10k.biom –metrics bray_curtis,unweighted_unifrac,weighted_unifrac
–t dada2.tre –o betaout
```

#### 7. Principal Coordinate Analysis (PCoA)

The Principal Coordinate Analysis (PCoA) is an ordination technique to summarize a beta diversity distance matrix into eigenvalues and eigenvectors. Often the principal coordinates are presented in two or three dimensions for ease of pattern identification (e.g., with a scatterplot). Given the weighted nature of NHANES data, PCoA vectors will need to be generated by the user taking into account the complex sampling design.

#### 8. Relative abundance/read count tables

The relative abundance and read count tables are available in the NCHS Research Data Center.

Relative abundance/read count tables were generated to characterize the percentage (relative) and the count of the bacterial reads in the sample. The relative abundance/read count tables were based on each phylogenetic level (i.e., 2: Phylum; 3: Class; 4: Order; 5: Family; 6: Genus) and conducted without rarefaction. The tables were generated using the summarize_taxa command in QIIME 1.9.1.

The relative abundance/read count tables were reformatted for each phylogenetic level and each method using an internal R script. Each row represents one human sample. Each column represents the database version and its corresponding phylogenetic level. For detailed taxonomy assignment for each phylogenetic level, an annotation file is provided for each pipeline.

QIIME1 Command logs:

summarize_taxa.py -i dada2_mod_final.biom -o taxa/relative -m mapping_human_all_noduplicate.txt -L 2 (2,3,4,5,6)
summarize_taxa.py -i dada2_mod_final.biom -o taxa/relative -m mapping_human_all_noduplicate.txt -L 2 -a (2,3,4,5,6)

#### Analytic Notes

The analysis of NHANES laboratory data must be conducted using the appropriate survey design and demographic variables. The NHANES 2009-2010 and NHANES 2011-2012 Demographics File contains demographic data, health indicators, and other related information collected during household interviews as well as the sample design variables. The recommended procedure for variance estimation requires use of stratum and PSU variables (SDMVSTRA and SDMVPSU, respectively) in the demographic data file.

This laboratory data file can be linked to the other NHANES data files using the unique survey participant identifier (i.e., SEQN).

Examination sample weights should be used for analyses. Please refer to the NHANES Analytic Guidelines and the on-line NHANES Tutorial for further details on the use of sample weights and other analytic issues.

#### Dictionary

| Term | Definition |
|---|---|
| 16S rRNA gene | A gene universally found in bacteria and archaea which is commonly used as a taxonomic marker gene, or a "genetic fingerprint," of different organisms. |
| Alpha diversity | Diversity within a single sample, typically representing microbial community richness and/or community evenness. Alpha diversity is commonly measured with metrics including observed OTUs/ASVs, Faith's Phylogenetic Diversity[10], the Shannon-Weiner index[11], and the Simpson index[12]. |
| Amplicon sequence variant (ASV) | A unique microbial sequence observed in a sample after quality control. In this data, ASVs are identified using DADA2 after error correction. |
| Beta diversity | Diversity between samples, typically representing pairwise dissimilarity of samples. Beta diversity is commonly measured between all pairs of samples with metrics including unweighted UniFrac, weighted UniFrac[13], and Bray-Curtis dissimilarity[14]. Measurement of beta diversity between all pairs of samples is typically represented in a distance matrix. |
| BIOM format | Biological observation matrix format (http://biom-format.org/), a community standard file format for feature tables[9]. |
| DADA2 pipeline | A bioinformatics tool that models and corrects sequencing errors in microbiome data to generate ASV feature tables which can resolve one nucleotide differences between sequences[6]. |

| | |
|---|---|
| Demultiplex | The process of associating 16S rRNA gene sequences with the sample that they were derived from based on DNA barcodes. |
| Feature table | A table that presents the read counts for each ASV or OTU for each individual. Typically, the file is stored in the BIOM format. |
| Operational taxonomic unit (OTU) | An undefined taxonomic grouping of organisms, intended to represent a concept such as a phylum, genus, or species. In some microbiome bioinformatics workflows, sequences are "clustered into OTUs," based on pairwise similarity to reduce the impact of erroneous sequence reads (by grouping them with sequences that do not contain errors), and reduce the computational burden of downstream bioinformatics steps (by reducing the number of sequences that need to be analyzed). |
| Phylogenetic tree | A representation of the hypothesized evolutionary relationships between ASVs, OTUs, or more generally, any biological entities. |
| Principal coordinate analysis (PCoA) | The Principal Coordinate Analysis (PCoA) is an ordination technique to summarize a beta diversity distance matrix into eigenvalues and eigenvectors. Often the principal coordinates are presented in two or three dimensions for ease of pattern identification (e.g., with a scatterplot). |
| QIIME | A bioinformatics tool designed to take users from raw microbiome sequencing data through publication quality graphics and statistics. This includes demultiplexing, quality filtering, ASV or OTU definition, taxonomic assignment, phylogenetic reconstruction, and diversity analyses and visualizations. QIIME is free for all use and open-source (https://qiime2.org)[5,8]. |
| Rarefaction | Random even subsampling of counts in a feature table without replacement to a specified total frequency. This is often applied to normalize the number of sequences observed across samples, as differential total frequencies can bias alpha and beta diversity metrics. |
| SILVA database | A 16S rRNA gene reference sequence database used to assign taxonomy[7]. |

**Flow Chart – Sample to datafiles**

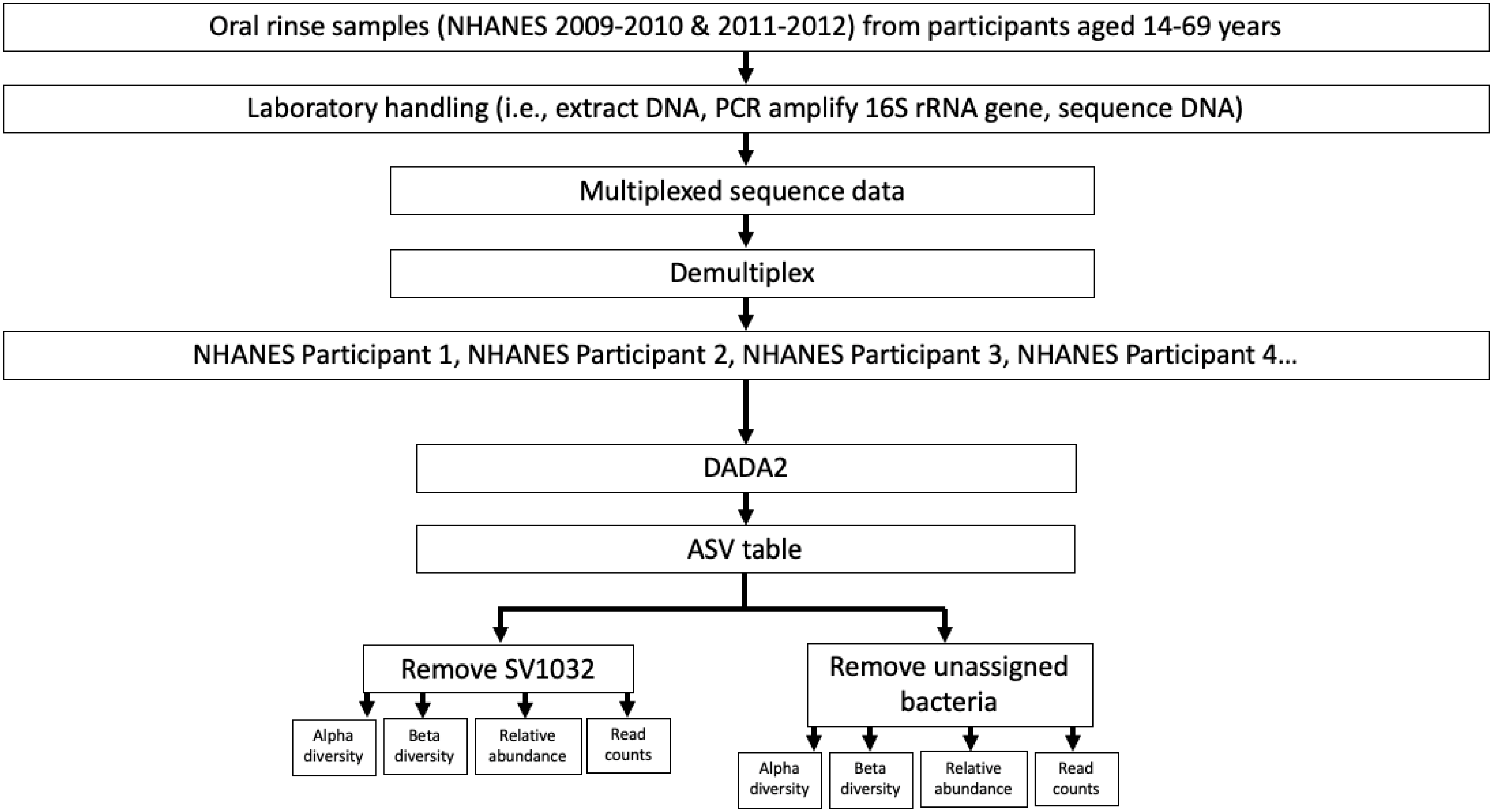

#### List of Files

| File Name | Data File | Variable/Annotation File | Access |
|---|---|---|---|
| ***Alpha Diversity*** | | | |
| Alpha Diversity - DADA2 (RB) | dada2rb-alpha.txt | | Public Use |
| Alpha Diversity variable list - DADA2 (RB) | | dada2rb-variablelist-alpha.txt | Public Use |
| Alpha Diversity - DADA2 (RSV) | dada2rsv-alpha.txt | | Public Use |
| Alpha Diversity variable list - DADA2 (RSV) | | dada2rsv-variablelist-alpha.txt | Public Use |
| ***Beta Diversity*** | | | |
| Beta Diversity Bray-Curtis - DADA2 (RB) | dada2rb-braycurtis-beta.txt | | Public Use |
| Beta Diversity unweighted Unifrac - DADA2 (RB) | dada2rb-unwunifrac-beta.txt | | Public Use |
| Beta Diversity weighted Unifrac - DADA2 (RB) | dada2rb-wunifrac-beta.txt | | Public Use |
| Beta Diversity Bray-Curtis - DADA2 (RSV) | dada2rsv-braycurtis-beta.txt | | Public Use |
| Beta Diversity unweighted Unifrac - DADA2 (RSV) | dada2rsv-unwunifrac-beta.txt | | Public Use |
| Beta Diversity weighted Unifrac - DADA2 (RSV) | dada2rsv-wunifrac-beta.txt | | Public Use |
| ***Relative Abundance/Read Count*** | | | |
| Read Count Phylum - DADA2 (RB) | dada2rb-phylum-count.txt | | Restricted |
| Relative Abundance Phylum - DADA2 (RB) | dada2rb-phylum_relative.txt | | Restricted |
| Read Count Class - DADA2 (RB) | dada2rb-class-count.txt | | Restricted |
| Relative Abundance Class - DADA2 (RB) | dada2rb-class-relative.txt | | Restricted |
| Read Count Order - DADA2 (RB) | dada2rb-order-count.txt | | Restricted |
| Relative Abundance Order - DADA2 (RB) | dada2rb-order-relative.txt | | Restricted |
| Read Count Family - DADA2 (RB) | dada2rb-family-count.txt | | Restricted |
| Relative Abundance Family - DADA2 (RB) | dada2rb-family-relative.txt | | Restricted |
| Read Count Genus - DADA2 (RB) | dada2rb-genus-count.txt | | Restricted |
| Relative Abundance Genus - DADA2 (RB) | dada2rb-genus-relative.txt | | Restricted |
| Taxonomy Annotation - DADA2 (RB) | | dada2rb-taxonomy-annotate.txt | Public Use |
| Read Count Phylum - DADA2 (RSV) | dada2rsv-phylum-count.txt | | Restricted |
| Relative Abundance Phylum - DADA2 (RSV) | dada2rsv-phylum-relative.txt | | Restricted |
| Read Count Class - DADA2 (RSV) | dada2rsv-class-count.txt | | Restricted |
| Relative Abundance Class - DADA2 (RSV) | dada2rsv-class-relative.txt | | Restricted |
| Read Count Order - DADA2 (RSV) | dada2rsv-order-count.txt | | Restricted |

| | | | |
|---|---|---|---|
| Relative Abundance Order - DADA2 (RSV) | dada2rsv-order-relative.txt | | Restricted |
| Read Count Family - DADA2 (RSV) | dada2rsv-family-count.txt | | Restricted |
| Relative Abundance Family - DADA2 (RSV) | dada2rsv-family-relative.txt | | Restricted |
| Read Count Genus - DADA2 (RSV) | dada2rsv-genus-count.txt | | Restricted |
| Relative Abundance Genus - DADA2 (RSV) | dada2rsvgenus_relative.txt | | Restricted |
| Taxonomy Annotation - DADA2 (RSV) | | dada2rsv-taxonomy-annotate.txt | Public Use |
