## Supplemental Table 7 for "The mouth of America: the oral microbiome profile of the US population"

**Table S7: Characteristics of NHANES 2009-2012 participants with or without oral microbiome data**

|  | **Without oral microbiome data, ages 18-69** | | **With oral microbiome data,**  **ages 18-69** | |
| --- | --- | --- | --- | --- |
|  | **N = 1861** | | **N = 8237** | |
|  | **N** | **(Weighted %) ^a^** | **N** | **(Weighted %) ^a^** |
| Age groups (years) |  |  |  |  |
| 18-29 | 409 | 21.6 | 2164 | 25.1 |
| 30-39 | 362 | 18.9 | 1573 | 19.6 |
| 40-49 | 368 | 20.6 | 1594 | 21.3 |
| 50-59 | 368 | 22.9 | 1445 | 20.0 |
| 60-69 | 354 | 16.1 | 1461 | 14.0 |
| Sex |  |  |  |  |
| Male | 1014 | 54.5 | 4127 | 50.0 |
| Female | 847 | 45.4 | 4110 | 50.0 |
| Self-reported race and ethnicity |  |  |  |  |
| Mexican American and other Hispanic | 476 | 13.6 | 2240 | 15.4 |
| Non-Hispanic Black | 790 | 68.6 | 3064 | 64.1 |
| Non-Hispanic White | 392 | 10.7 | 196 | 12.4 |
| Non-Hispanic other race including multiracial | 203 | 7.2 | 972 | 8.0 |
| Education |  |  |  |  |
| Less than high school | 490 | 1.1 | 2053 | 17.0 |
| Completed high school/GED | 422 | 22.5 | 1831 | 21.1 |
| More than high school | 949 | 60.4 | 4353 | 61.9 |
| Marital status |  |  |  |  |
| Never married | 473 | 23.2 | 2267 | 24.4 |
| Married/living with partner | 1029 | 60.3 | 4560 | 60.7 |
| Divorced/widowed/separated | 359 | 16.5 | 1410 | 14.9 |
| Income-to-poverty ratio |  |  |  |  |
| <1 (below poverty level) | 434 | 15.0 | 1985 | 16.7 |
| 1-1.999 | 409 | 17.2 | 1975 | 18.4 |
| 2-2.999 | 237 | 13.9 | 947 | 12.7 |
| ≥3 | 570 | 46.3 | 2641 | 45.4 |
| Missing | 211 | 7.6 | 689 | 6.7 |
| Body mass index (kg/m^2^) categories |  |  |  |  |
| <18.5 (Underweight) | 47 | 2.4 | 150 | 1.9 |
| 18.5-24.999 (Normal weight) | 595 | 31.5 | 2463 | 30.5 |
| 25-29.999 (Overweight) | 563 | 32.8 | 2614 | 32.4 |
| 30-34.999 (Obesity) | 377 | 18.9 | 1653 | 20.1 |
| ≥35 (Severe obesity) | 279 | 14.3 | 1357 | 15.1 |
| Cigarette smoking history |  |  |  |  |
| Never | 1058 | 56.9 | 4051 | 56.2 |
| Former | 311 | 18.4 | 1408 | 18.2 |
| Current | 492 | 24.7 | 2178 | 25.5 |
| Alcohol consumption |  |  |  |  |
| Never drinker | 213 | 7.9 | 1032 | 9.3 |
| Drinks 0 drinks/week in past 12 months | 191 | 8.9 | 1124 | 12.3 |
| Drinks >0-<1 drink/week in past 12 months | 382 | 21.5 | 2351 | 28.0 |
| Drinks 1-<8 drinks/week in past 12 months | 387 | 24.9 | 1931 | 27.7 |
| Drinks 8-<14 drinks/week in past 12 months | 77 | 5.0 | 527 | 7.9 |
| Drinks ≥14 drinks/week in past 12 months | 102 | 6.2 | 559 | 7.5 |
| Missing | 509 | 25.6 | 713 | 7.4 |
| Diabetes |  |  |  |  |
| No | 1624 | 89.6 | 7210 | 90.1 |
| Yes | 237 | 10.4 | 1117 | 9.9 |
| Hypertension |  |  |  |  |
| No | 1063 | 59.1 | 1861 | 58.2 |
| Yes | 798 | 40.9 | 4621 | 41.8 |
| Periodontal disease/edentulism |  |  |  |  |
| None | 365 | 25.3 | 2551 | 37.8 |
| Mild | 57 | 2.7 | 365 | 4.3 |
| Moderate | 252 | 13.0 | 1720 | 18.4 |
| Severe | 84 | 4.1 | 676 | 5.8 |
| Not eligible (<30 years old) | 409 | 21.6 | 2164 | 25.1 |
| Edentulous | 47 | 1.6 | 271 | 2.5 |
| Missing | 647 | 31.8 | 490 | 6.0 |
| Antibiotic use |  |  |  |  |
| No | 1796 | 96.3 | 7949 | 96.2 |
| Yes | 65 | 3.7 | 288 | 3.8 |
| Anti-gastroesophageal reflux medication |  |  |  |  |
| No | 1684 | 89.5 | 1861 | 91.6 |
| Yes | 177 | 10.5 | 7537 | 8.5 |
| Anti-hyperlipidemic medication |  |  |  |  |
| No | 1618 | 86.6 | 7158 | 86.8 |
| Yes | 243 | 13.4 | 1079 | 13.2 |
| Inhaled respiratory medication |  |  |  |  |
| No | 1851 | 99.6 | 8186 | 99.4 |
| Yes | 10 | 0.4 | 51 | 0.6 |

^a^ Weighted estimates derived using NHANES MEC weights
