## Supplemental Table 13 for "The mouth of America: the oral microbiome profile of the US population"

**Table S13: Variable definitions and code**

| **Code used for variable creation** | 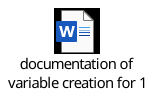 |
| --- | --- |
| **Variable** | **Source/creation** |
| **Age**, modeled as restricted 5-knot cubic regression splines | Based on self-reported age at interview |
| **Sex** | Based on self-reported sex at interview |
| Male |  |
| Female |  |
| **Self-reported race and ethnicity** | Based on self-reported race and ethnicity at interview. No collapsing of groups was conducted. |
| Mexican American and other Hispanic |  |
| Non-Hispanic Black |  |
| Non-Hispanic White |  |
| Non-Hispanic other race including multiracial |  |
| **Education** | Self-reported level of highest education. Missing values were assigned the mode—more than high school. |
| Less than high school |  |
| Completed high school/GED |  |
| More than high school |  |
| **Marital status** | Self-reported marital status. Missing values were assigned the mode—married/living with partner. |
| Never married |  |
| Married/living with partner |  |
| Divorced/widowed/separated |  |
| **Income-to-poverty ratio** | Self-reported family income divided by the federal poverty level. Missing values were coded as a separate category. |
| <1 (below poverty level) |  |
| 1-1.999 |  |
| 2-2.999 |  |
| ≥3 |  |
| Missing |  |
| **Body mass index (kg/m^2^) categories** | Based on measured height and weight. Missing values were assigned the mode—normal weight category. |
| <18.5 (Underweight) |  |
| 18.5-24.999 (Normal weight) |  |
| 25-29.999 (Overweight) |  |
| 30-34.999 (Obesity) |  |
| ≥35 (Severe obesity) |  |
| **Cigarette smoking history** | Self-reported smoking status, does not consider serum cotinine levels. Missing values were assigned the mode—never smoker. |
| Never |  |
| Former |  |
| Current |  |
| **Alcohol consumption** | Self-reported alcohol intake over the past 12 months, expressed as drinks per week. Missing values were coded as a separate category. |
| Never drinker |  |
| Drinks 0 drinks/week in past 12 months |  |
| Drinks >0-<1 drink/week in past 12 months |  |
| Drinks 1-<8 drinks/week in past 12 months |  |
| Drinks 8-<14 drinks/week in past 12 months |  |
| Drinks ≥14 drinks/week in past 12 months |  |
| Missing |  |
| **Diabetes** | Based on self-report of diabetes, self-report of taking insulin or other diabetes medication, measured hemoglobin A1C of >= 6.5, or measured serum glucose levels >=126. Missing values were assigned the mode—no diabetes. |
| No |  |
| Yes |  |
| **Hypertension** | Based on self-report of hypertension, self-report of taking hypertension medication, measured diastolic pressure >80 or measured systolic pressure >130. Missing values were assigned the mode—no hypertension. |
| No |  |
| Yes |  |
| **Periodontal disease/edentulism** | Combined variable for severity of periodontal disease and edentulism. Periodontal disease severity was based on periodontal disease examination; edentulism was based on measured count of teeth. Missing values were coded as a separate category. |
| None |  |
| Mild |  |
| Moderate |  |
| Severe |  |
| Not eligible (<30 years old) |  |
| Edentulous |  |
| Missing |  |
| **Antibiotic use** | Based on self-reported use of medications in the past 30 days. Missing values were assigned the mode—no use. |
| No |  |
| Yes |  |
| **Anti-gastroesophageal reflux medication** | Based on self-reported use of medications in the past 30 days. Missing values were assigned the mode—no use. |
| No |  |
| Yes |  |
| **Anti-hyperlipidemic medication** | Based on self-reported use of medications in the past 30 days. Missing values were assigned the mode—no use. |
| No |  |
| Yes |  |
| **Inhaled respiratory medication** | Based on self-reported use of medications in the past 30 days. Missing values were assigned the mode—no use. |
| No |  |
| Yes |  |

**NOTE**: Missing values were assigned to the most prevalent category (mode) when fewer than 5% of participants had missing observations. Missing values were treated as a separate category when >5% of participants had missing observations.
