## Supplementary figures and images for "The mouth of America: the oral microbiome profile of the US population"

### Supplemental figure 3

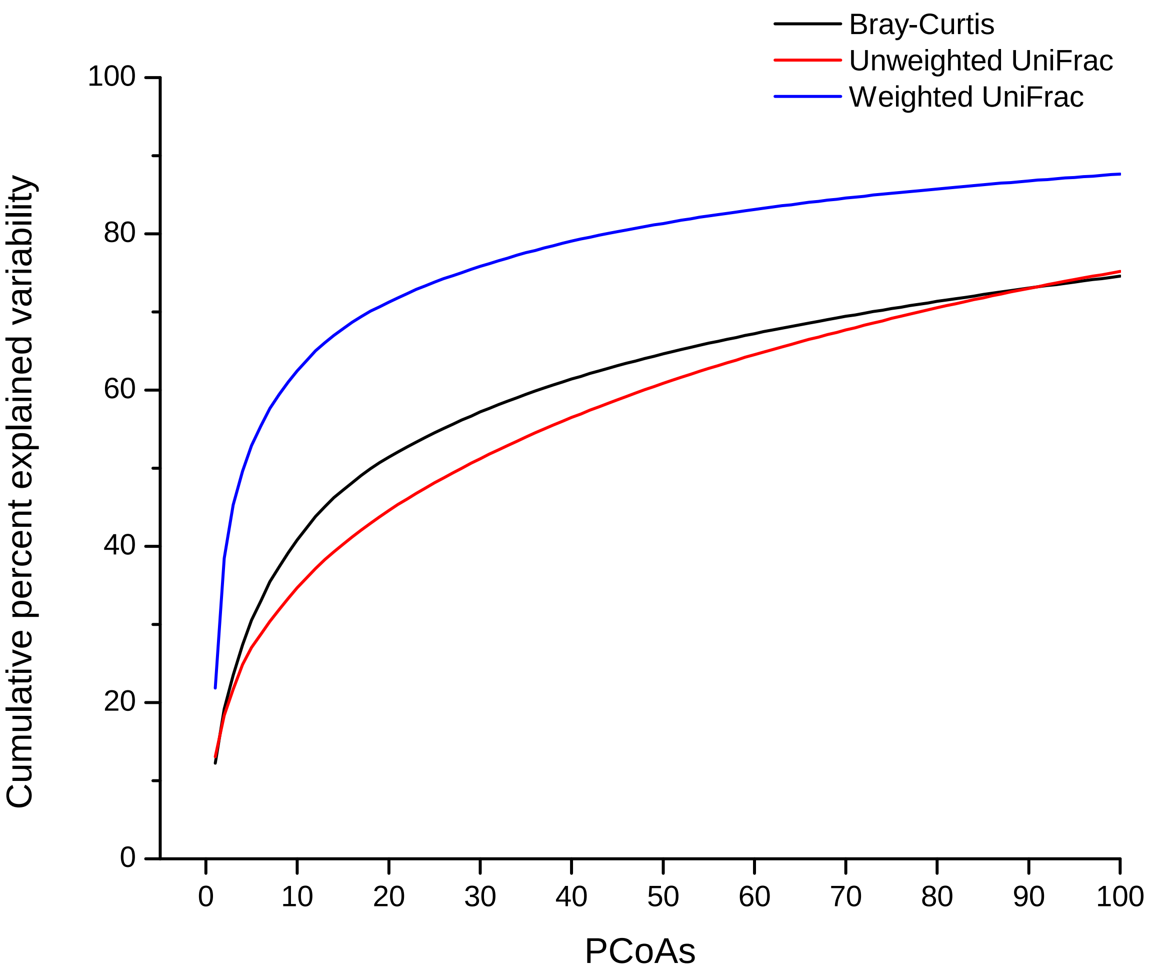

### Supplemental figure 7

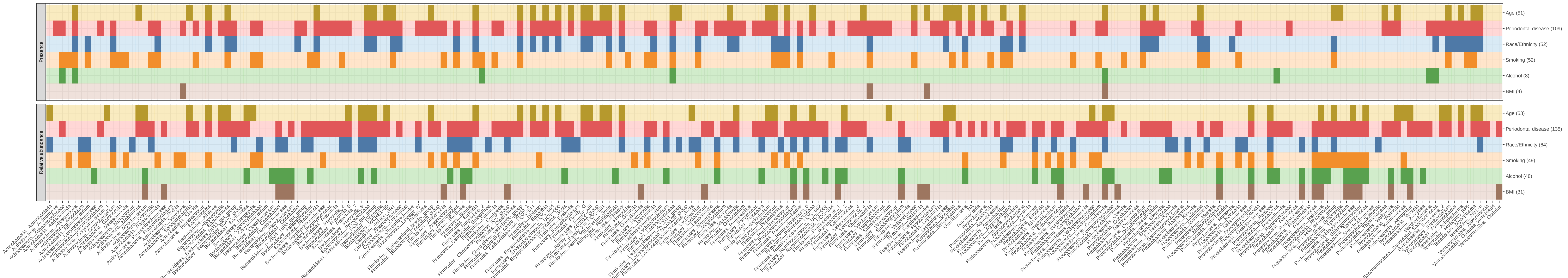

### Supplemental figures 1 & 2

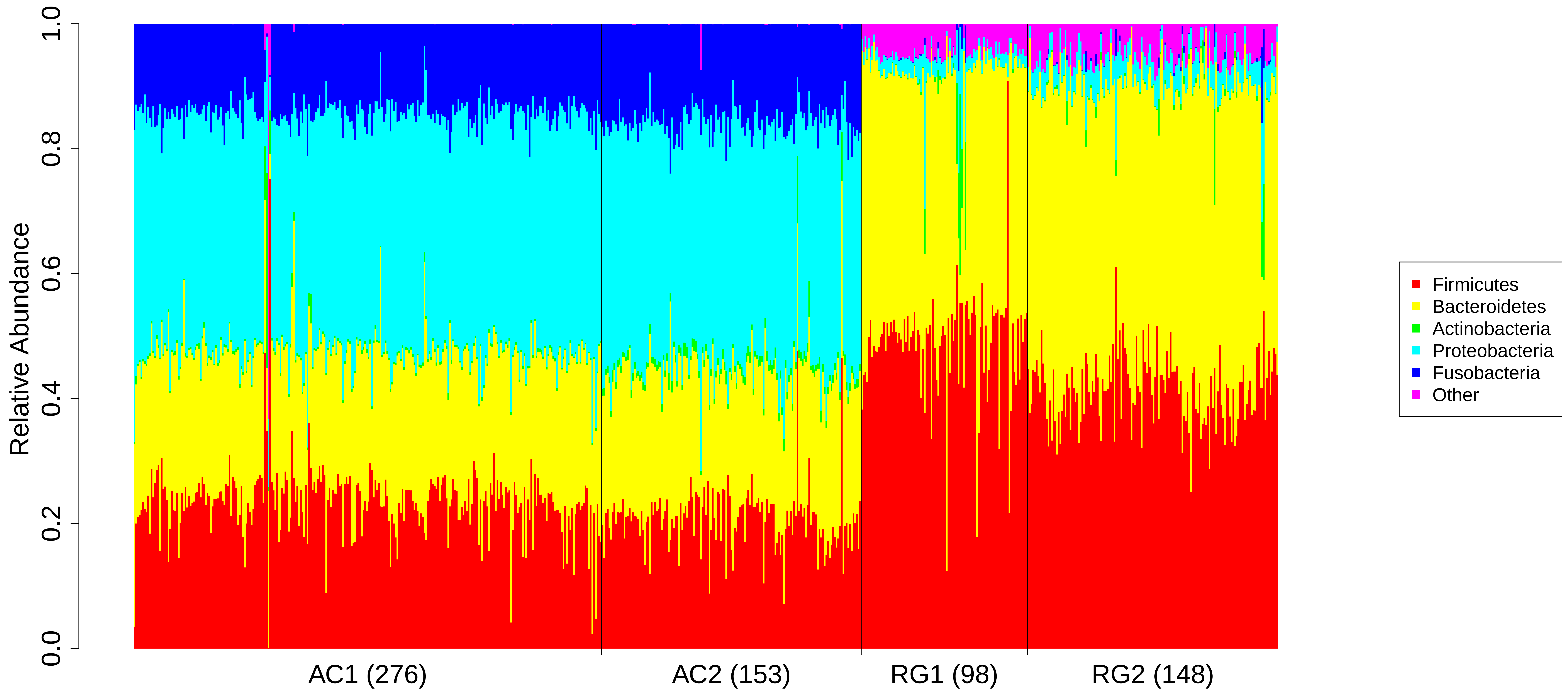

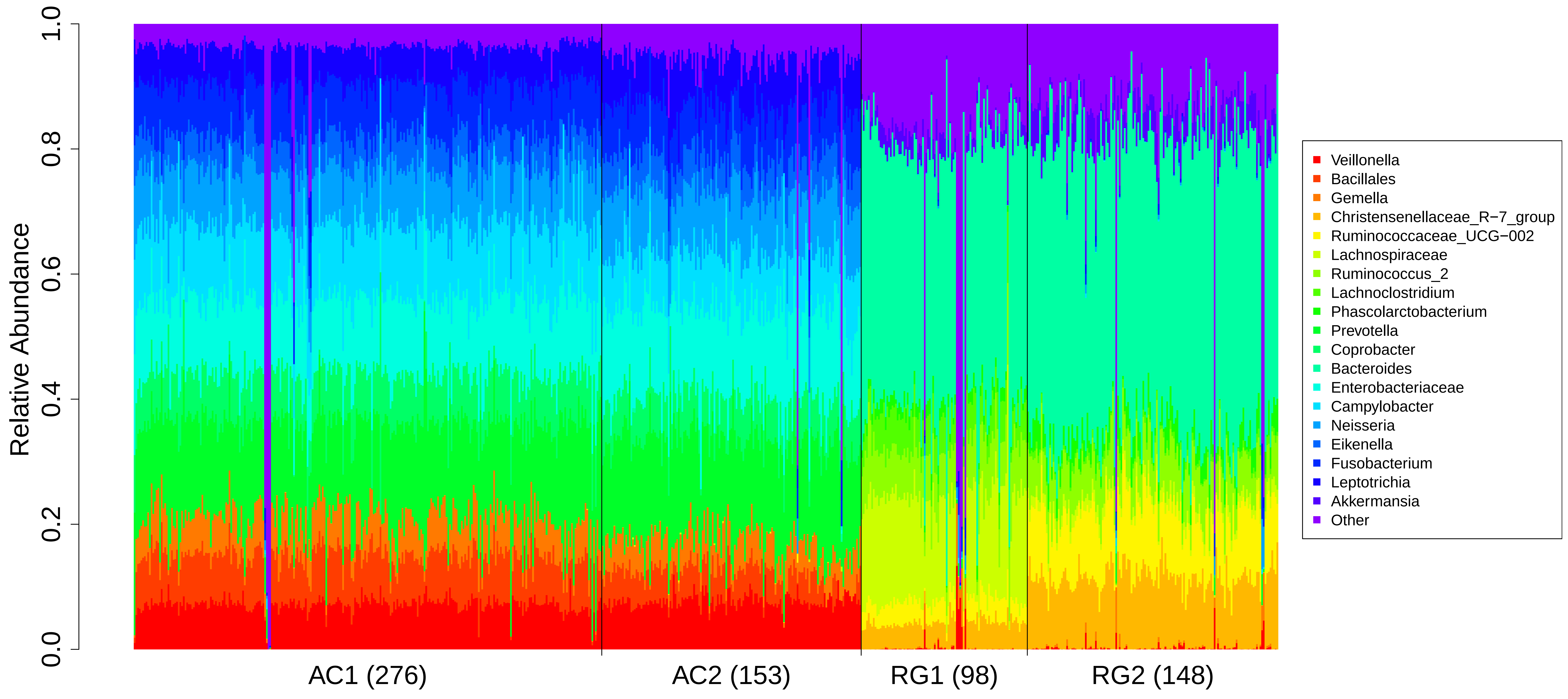

### Supplemental figures 4-6

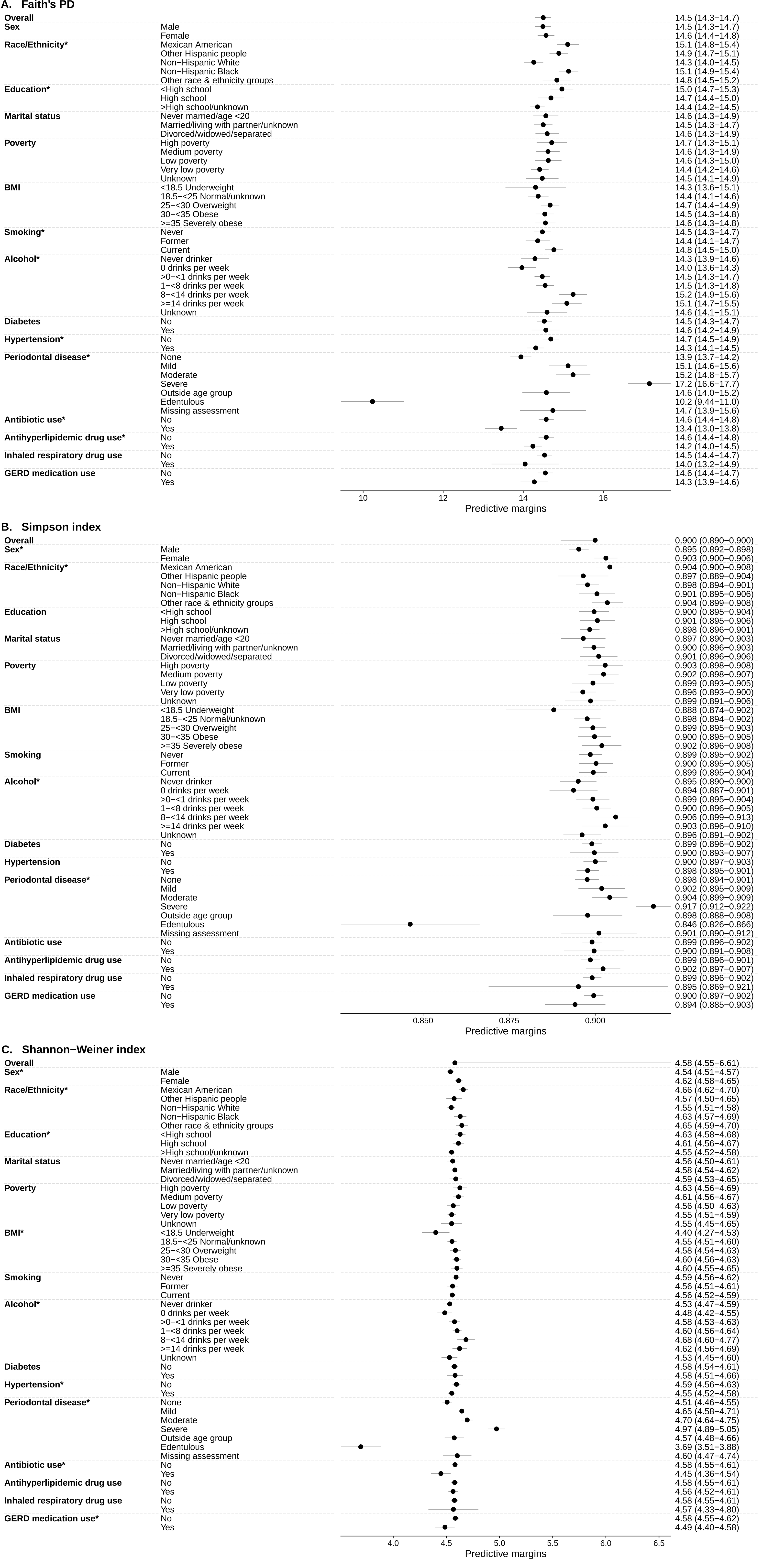
